# Long COVID affects working memory: Assessment using a single rapid online test

**DOI:** 10.1101/2025.01.26.25321152

**Authors:** Aziz UR Asghar, Hong Kiu Yuen, Murat Aksoy, Abayomi Salawu, Heidi A Baseler

## Abstract

Long COVID, or post-COVID-19 syndrome, is characterized by persistent symptoms following SARS-CoV-2 infection, including cognitive impairments such as brain fog that adversely affects quality of life. The aim of this study was to evaluate the impact of long COVID on working memory using a single rapid online anonymous survey and working memory quiz. We analyzed working memory scores in relation to long COVID status (clinically diagnosed, self-reported, and non-long COVID), age, number of COVID-19 infections and long COVID duration, and subjective ratings of brain fog severity and overall life impact of symptoms. The study recruited 1064 participants aged 16 to over 85 years (83% from the United Kingdom) with 39% reporting long COVID. The results demonstrate that long COVID significantly impairs working memory, with the greatest deficits observed in clinically diagnosed cases. In addition, we found that working memory declined with age and multiple COVID-19 infections particularly in the diagnosed long COVID group. Moreover, individuals in the diagnosed long COVID group reported experiencing the most severe brain fog and the greatest impact of long COVID symptoms on their lives, and both of these factors were strongly correlated with lower memory scores. Participants who had been living with long COVID for a longer duration had lower memory scores. However, this relationship was likely influenced by the SARS-CoV-2 variant, as individuals with prolonged long COVID symptoms were more likely to have been infected with earlier variants (alpha and wild-type) which are linked to more severe disease outcomes. Our results suggest that long COVID is associated with significant working memory impairments, emphasizing the need for targeted interventions and support strategies for those with long COVID to address memory problems.

## Introduction

Long COVID, also known as post COVID-19 syndrome/condition, broadly refers to the signs and symptoms that develop during or after an infection with the SARS-CoV-2 virus that persists over a number of months and has no alternative explanation (NICE, 2020; WHO, 2022). A systematic review of long COVID prevalence, which included 194 studies with 735,006 participants, found that 45% of those who survived COVID-19 continued to experience symptoms at approximately four months (O’Mahoney et al., 2023).

Approximately 1.9 million people in the United Kingdom self-reported long COVID symptoms with 41% experiencing symptoms for at least 2 years and 79% reported that their symptoms negatively impacted their daily activities (ONS, 2023); over 200 long COVID symptoms have been documented, and the most commonly reported were fatigue (72%), difficulty concentrating (51%), muscle aches (49%) and shortness of breath (48%).

Substantial cognitive performance deficits in tasks requiring memory, attention and reasoning have been reported for people with COVID-19 symptoms that persisted for up to two years (Bertuccelli et al., 2022; Cheetham et al., 2023; Kim et al., 2023; Millet et al., 2022; Zawilska & Kuczynska, 2022; Zhao et al., 2023; Ziauddeen et al., 2022). The cognitive impact of long COVID, often generally described as ‘brain fog’ (including problems with memory and performing daily tasks), is a major contributor to poor functioning and reduced quality of life experienced by patients with long COVID (Jennings et al., 2022; Lam et al., 2023; Nalbandian et al., 2021; Nordvig et al., 2023). A recent systematic review and meta-analysis of studies evaluating brain fog, cognitive function and mental health symptoms in people with long COVID found that the average overall prevalence of memory loss was 21% and as high as 61% (van der Feltz-Cornelis et al., 2024). This considerable variation in memory loss prevalence likely reflects differences in how memory was defined, assessed or self-reported across studies.

Working memory is the process of temporarily storing and retrieving information relevant to performing the current task (Baddeley et al., 2019; Repovs & Baddeley, 2006). Working memory is integral in everyday tasks including problem solving, holding a conversation, reading comprehension and decision making. Working memory deficits have been consistently reported when testing cognitive function in long COVID (Graham et al., 2021; Guo et al., 2022; Miskowiak et al., 2021; Miskowiak et al., 2023).

Studies investigating working memory deficits related to COVID-19 or long COVID are often measured within the context of a battery of multiple cognitive tests (Arbula et al., 2024; Cheetham et al., 2023; Cui et al., 2024; Graham et al., 2021; Guo et al., 2022; Miskowiak et al., 2021; Miskowiak et al., 2023; Stavem et al., 2022). This is potentially problematic for two reasons. Firstly, when embedded within a more complex cognitive task (for example, spatial mental rotation), it is difficult to interpret performance on the working memory component alone. Moreover, it is possible that a participant’s performance on a working memory task could be affected by previous different cognitive tasks. Secondly, undertaking numerous cognitive tests within a given study requires statistical correction for multiple comparisons, which reduces the statistical power and increases the likelihood of type II errors (false negatives). Conversely, if statistical correction is not applied to correct for multiple cognitive tests, there is the risk for type I errors (false positives). Since the majority of people with long COVID report fatigue and difficulty in concentrating as major symptoms, completing a lengthy survey with multiple cognitive tests would be burdensome and may decrease completion rates or yield unreliable data. We have previously shown, using a single cognitive task requiring minimal time and effort, that working memory was negatively impacted in people who had been infected with COVID-19 compared to those who reported not having had COVID-19 (Baseler et al., 2022).

The main aim of our study was to investigate the impact of long COVID on working memory function. To capture a broad demographic sample, we distributed an online survey with a single working memory quiz that could be completed relatively rapidly. We utilized the same memory quiz as we have used previously to demonstrate the negative impact of COVID-19 on working memory function, which was anonymous and incorporated elements of gamification to encourage more people to fully complete the survey and memory quiz (Baseler et al., 2022). In our analysis we evaluate the impact of several variables on objective memory scores including long COVID status (diagnosed long COVID, self-reported long COVID and non-long COVID), age, gender, number of COVID-19 infections and long COVID duration. In addition, we examined the relationship between objective memory scores and participant subjective ratings of brain fog, and the overall impact of long COVID symptoms on their life.

## Results

### Demographics of participants

We included 1064 participants that fully completed the L-CORONA survey and memory quiz in the period between 16/03/2023 and 31/12/2023. Participant demographics are summarized in Table 1. In terms of long COVID status, 39% of participants had long COVID (N=415), and 61% did not have long COVID (‘Non-Long COVID group’). Participants ranged from 16 to over 85 years of age, with the highest number of participants in the age range 45 to 54 years old. The majority of the participants resided in the United Kingdom (83%), with the remainder residing in 25 other countries around the globe. Over 94% of participants (N=1001) completed the survey and memory quiz within 15 minutes (median = 7.2 minutes): long COVID group (median = 7.3 mins, N=393); non-long COVID group (median = 6.9 minutes, N=608).

**Table 1.** Participant demographics.

| **Long COVID status** | **N** | **%** |
| --- | --- | --- |
| Non-Long COVID | 649 | 61.0 |
| Long COVID | 415 | 39.0 |
| **Age (Years)** |  |  |
| 16 - 17 | 14 | 1.3 |
| 18 - 24 | 128 | 12.0 |
| 25 - 34 | 184 | 17.3 |
| 35 - 44 | 223 | 21.0 |
| 45 - 54 | 262 | 24.6 |
| 55 - 64 | 188 | 17.7 |
| 65 - 74 | 49 | 4.6 |
| 75 - 84 | 15 | 1.4 |
| 85+ | 1 | <1.0 |
| **Gender** |  |  |
| Female | 806 | 75.8 |
| Male | 244 | 22.9 |
| Other | 14 | 1.3 |
| **Countries** |  |  |
| UK | 883 | 83.0 |
| Non-UK | 181 | 17.0 |

### Categorical regression and objective memory scores

A categorical regression was used to assess the relative contributions of long COVID status (non-long COVID, diagnosed long COVID, or self-reported long COVID), age, number of COVID-19 infections and gender to memory scores in all participants.

Table 2 shows that significant effects were found (all *p*-values < 0.001), in order of importance, for long COVID status (55.4%), followed by age (31.5%) and number of COVID-19 infections (13.3%). Gender did not have a significant effect on memory scores (*p* > 0.05).

**Table 2.** Categorical regression analysis and objective memory scores for non-long COVID, diagnosed long COVID and self-reported long COVID participants.

| Adjusted regression coefficient $R^2 = 0.114$, Fisher's $F$-test = 13.479, $p < 0.001$ | | | | |
| --- | --- | --- | --- | --- |
| Independent Variables | Beta | $F$ | $p$ | Importance (Pratt) |
| Long COVID status<br>(non-long COVID, diagnosed long COVID, self-reported long COVID groups) | 0.240 | 60.221 | **< 0.001** | 0.554 |
| Age | -0.184 | 48.074 | **< 0.001** | 0.315 |
| Number of COVID-19 infections | -0.110 | 7.686 | **< 0.001** | 0.133 |
| Gender | 0.011 | 0.402 | 0.669 | -0.002 |

Significant $p$ -values are given in bold.

A second categorical regression was performed for the two long COVID groups only (those who self-reported having long COVID or were diagnosed with long COVID) to assess the additional contribution of long COVID duration on memory scores. Table 3 shows that the significant factors were, in order of importance: age (52.9%, *p* < 0.001), long COVID duration (26.0%, *p* < 0.001), and number of COVID-19 infections (15.5%, *p* = 0.003). Long COVID group (self-reported *versus* diagnosed long COVID) or gender did not have a significant effect on memory scores (*p* > 0.05).

**Table 3.** Categorical regression analysis and objective memory scores for diagnosed long COVID and self-reported long COVID participants only.

| Adjusted regression coefficient $R^2 = 0.079$ Fisher's $F$-test= 3.207, $p < 0.001$ | | | | |
| --- | --- | --- | --- | --- |
| Independent Variables | Beta | $F$ | $p$ | Importance (Pratt) |
| Age | -0.238 | 32.297 | **<0.001** | 0.529 |
| Long COVID duration | -0.145 | 6.491 | **<0.001** | 0.260 |
| Number of COVID infections | -0.139 | 4.678 | **0.003** | 0.155 |
| Long COVID groups only (diagnosed long COVID, self-reported long COVID groups) | 0.044 | 1.360 | 0.244 | 0.055 |
| Gender | 0.007 | 0.044 | 0.957 | 0.001 |

Significant $p$ -values are given in bold.

Based on these categorical regressions (Tables 2 and 3), we selected the following main factors affecting objective memory scores and evaluated each of them independently: long COVID status, age, number of COVID-19 infections, and for the long COVID groups only long COVID duration.

### Effect of long COVID status on working memory

Fig 1a shows that the objective memory scores are significantly reduced in the two long COVID groups combined (Mean (M) ± standard error of the mean = 14.15 ± 0.10) compared to the non-long COVID group (M = 15.02 ± 0.05, Mann-Whitney U = 167938.50, Z = −7.12, *p* < 0.001, r = 0.22). Next, we divided the long COVID group into sub-groups: those who suspected they may have long COVID (self-reported), and those who were diagnosed with long COVID by a medical professional. Fig 1b shows that objective memory scores were highest in the non-long COVID group but were significantly reduced in each of the long COVID sub-groups, with the lowest scores in the group diagnosed with long COVID. A Kruskal-Wallis test revealed a significant difference between the mean memory scores across groups [H(2) = 61.38, *p* < 0.001]. Dunn’s pairwise comparison tests with Bonferroni correction revealed significant differences between the non-long COVID group (M = 15.02 ± 0.05) and the self-reported long COVID (M = 14.48 ± 0.14, *p* = 0.0042), and between the non-long COVID and diagnosed long COVID (M = 13.88 ± 0.15, *p* < 0.001) groups. In addition, there was a significant difference between the self-reported long COVID (M = 14.48 ± 0.14) and diagnosed (M = 13.88 ± 0.15, *p* = 0.0031) long COVID groups.

**Fig 1.**
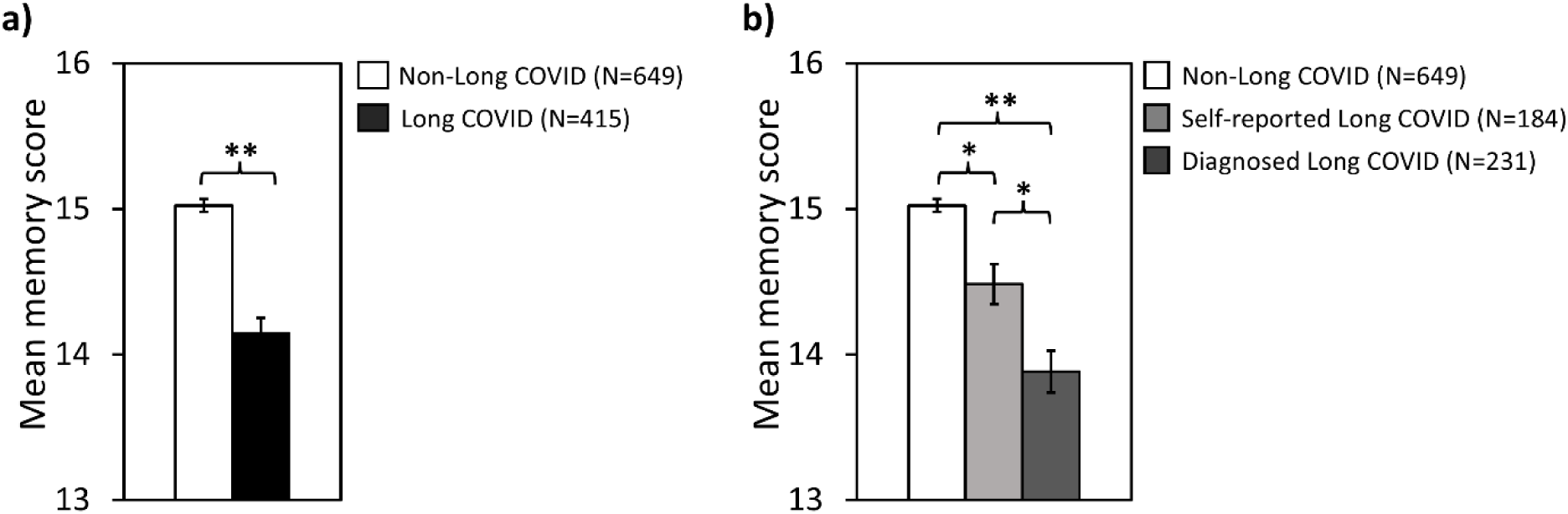
The effect of long COVID status on objective memory scores. **a)** Non-long COVID group compared to both long COVID groups combined (diagnosed plus self-reported long COVID), and **b)** Non-long COVID group compared to self-reported and diagnosed long COVID groups separately. Error bars represent standard error of the mean. \**p* < 0.01, \*\**p* < 0.001.

### Effect of age on working memory

We found that objective memory scores decreased significantly with age for both the non-long COVID and the diagnosed long COVID groups but not for the self-reported long COVID group. A Kendall’s tau-b correlation showed a negative relationship between mean memory score and age for both the non-long COVID (τb = –0.155, *p* < 0.001) and diagnosed long COVID (τb = –0.208, *p* < 0.001) groups across the nine age categories. The self-reported long COVID group did not have a significant relationship between age and mean memory scores (τb = –0.099, *p* = 0.098).

Participants aged 16-17 years old were combined with those aged 18 to 24 years old, and all participants above 55 years old were combined into a single group to increase sample size due to the lower numbers of participants who completed the memory quiz in these age categories (see Table 1). Fig 2 shows the effect of age on memory scores for all three groups for the five age categories. A Kruskal-Wallis test was performed at each of the five age categories to determine differences between groups based on long COVID status. There was no significant effect of long COVID status on memory scores for the 16-24 year-old age category [H(2) = 4.201, *p* = 0.122], or the 25-34 year-old age category [H(2) = 0.004, *p* = 0.998]. However, there was a significant effect of long COVID status on memory scores for the 35-44 [H(2) = 18.539, *p* < 0.001], 45-54 [H(2) = 24.377, *p* < 0.001] and 55+ year-old categories [H(2) = 15.416, *p* < 0.001]. Dunn’s pairwise comparisons (with Bonferroni correction for multiple tests) revealed that the memory scores in the diagnosed long COVID group were significantly reduced compared to the non-long COVID group in the 35-44, 45-54 and 55+ year-old categories (*p* < 0.001). The memory scores for the self-reported long COVID group were significantly reduced compared to the non-long COVID group in the 35-44 age category (*p* = 0.021). In the 55+ age category, the memory scores for the diagnosed long COVID group were significantly smaller than for the self-reported long COVID group (*p* = 0.034).

**Fig 2.**
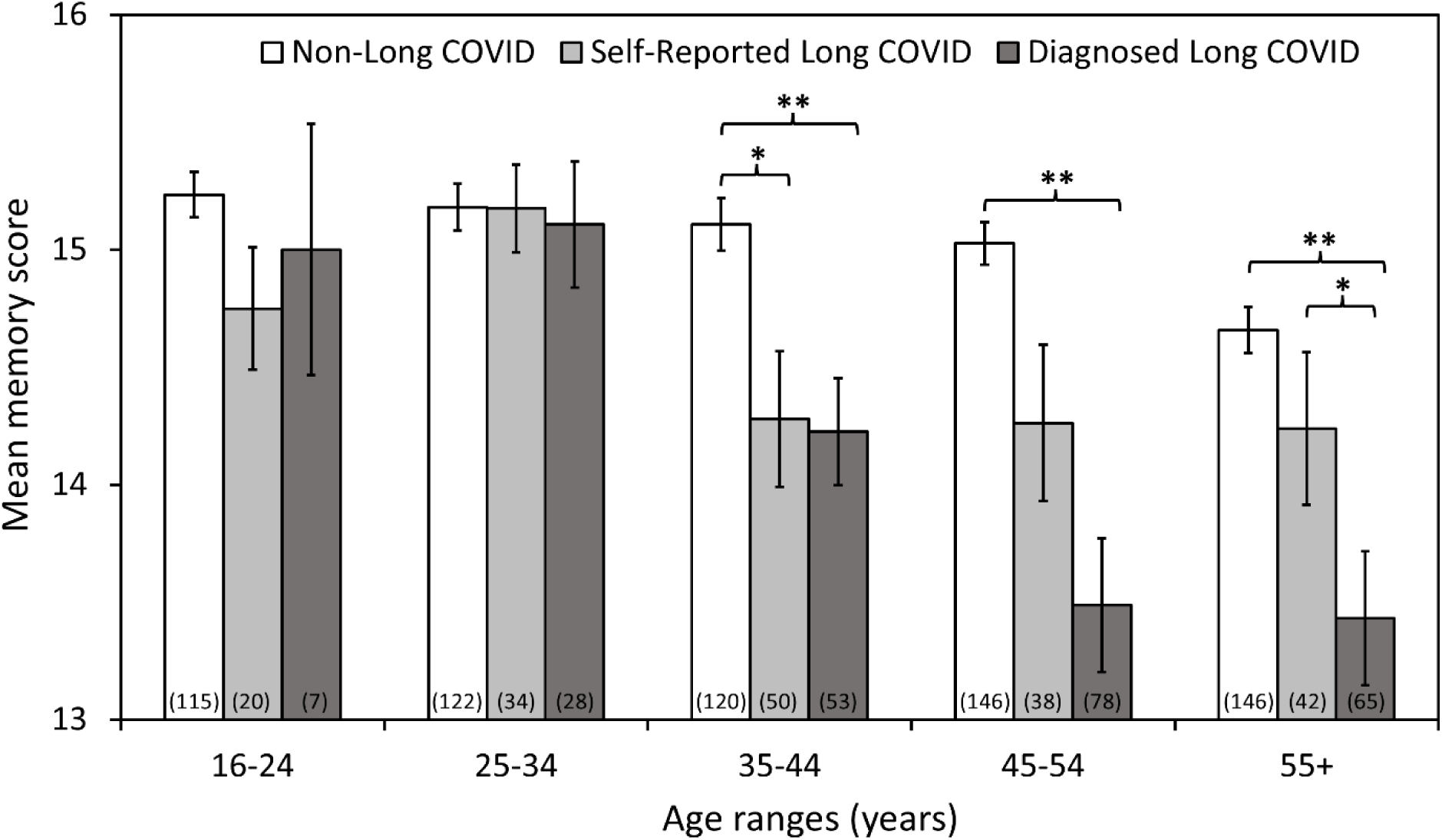
The effect of age on objective memory scores in the non-long COVID, self-reported long COVID and diagnosed long COVID groups. The sample size is given in brackets for each data bar. Error bars represent the standard error of the mean. Asterisks indicate significant differences between groups for each age range category. \**p* < 0.05, \*\**p* < 0.001.

### Effect of the number of COVID-19 infections on working memory

Objective memory scores decreased significantly with increasing number of COVID-19 infections for the diagnosed long COVID group (τb = –0.113, *p* = 0.043). However, there was no significant correlation between the number of COVID-19 infections and memory scores for either the self-reported long COVID group (τb = –0.040, *p* = 0.527) or the non-long COVID group (τb = 0.001, *p* = 0.970). Next, we compared the impact of the number of COVID-19 infections on memory scores between the non-long COVID, self-reported long COVID and diagnosed long COVID groups. Since there were only 13 participants with four or more COVID-19 infections we combined these participants with those who had three COVID-19 infections (N=67) to give a total number of 80 participants in this category. Fig 3 shows the effect of the number of COVID-19 infections on memory scores for all three long COVID status groups for the four infection categories. A Kruskal-Wallis test was performed at each of the four COVID-19 infection categories to determine differences between groups based on long COVID status. When participants had one or two COVID-19 infections more than 12 weeks prior to completing the survey and memory quiz, there was a significant effect of long COVID status on memory scores [one COVID-19 infection: H(2) = 23.046, *p* < 0.001; two COVID-19 infections: H(2) = 25.352, *p* < 0.001]. There was no significant effect of long COVID status on memory scores in participants with 3 or more infections [H(2) = 5.228, *p* = 0.073]. Dunn’s pairwise comparisons (with Bonferroni correction for multiple tests) revealed that the memory scores in the diagnosed long COVID group were significantly reduced compared to the non-long COVID group in the one and two COVID-19 infection categories (*p* < 0.001). The memory scores for the diagnosed long COVID group were significantly reduced compared to the self-reported long COVID group in the two COVID-19 infection category (*p* = 0.007). For the zero number of COVID-19 infection category, there was an insufficient number of participants in the self-reported (N=5) and diagnosed long COVID (N=1) groups to perform statistical analysis (these participants stated having long COVID despite an apparent absence of COVID-19 infection).

**Fig 3.**
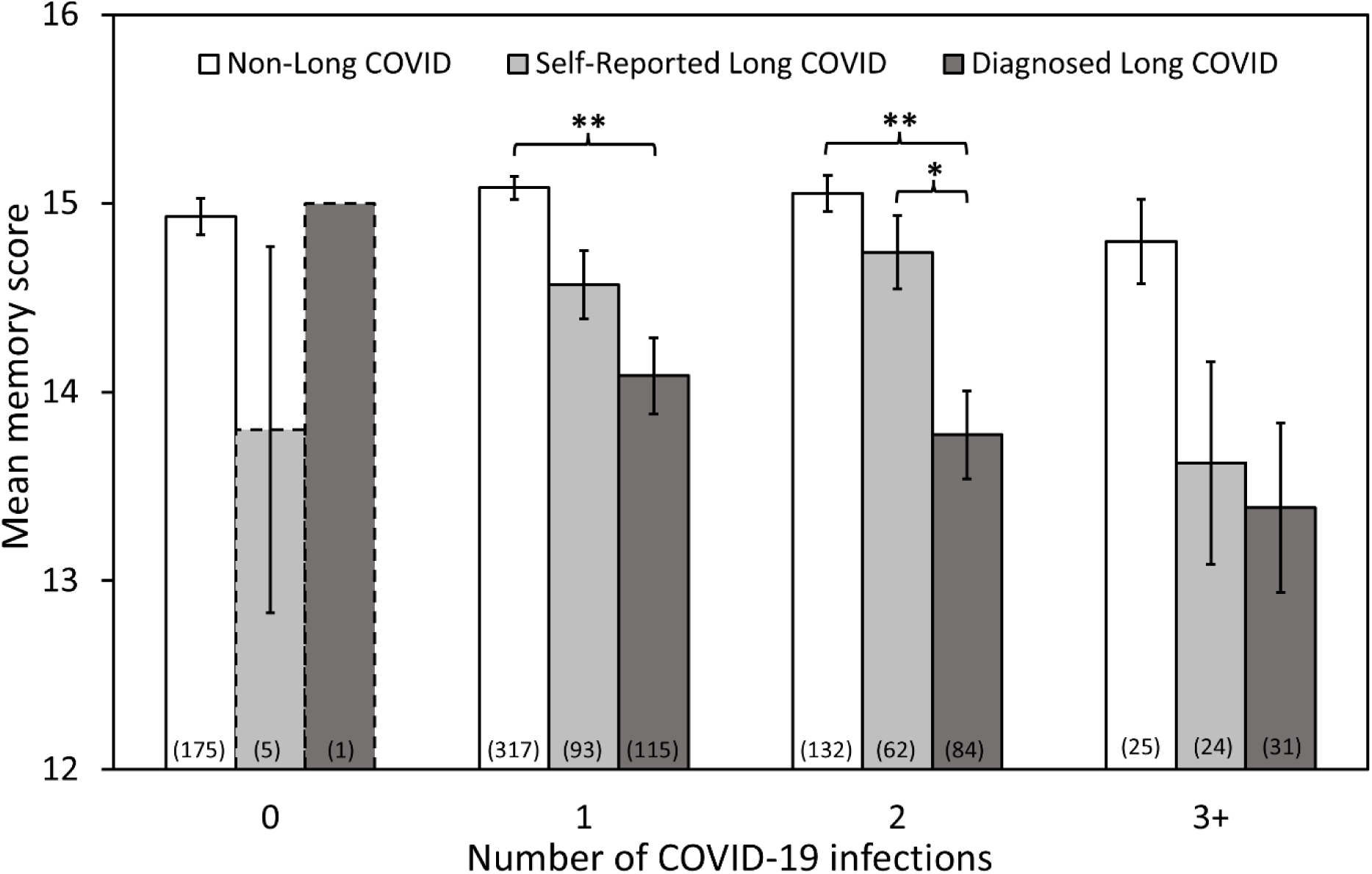
The effect of COVID-19 infections on objective memory scores in the non-long COVID, self-reported long COVID and diagnosed long COVID groups. The sample size is given in brackets for each data bar. Error bars represent the standard error of the mean. Asterisks indicate significant differences between groups for the number of COVID-19 infections. The dashed border on vertical bars (zero COVID-19 infections category) indicates the small number of participants who stated having long COVID despite an apparent absence of COVID-19 infection. \**p* < 0.01, \*\**p* < 0.001

### Effect of long COVID duration on working memory

As the number of months of having long COVID increased, the memory scores decreased significantly in the diagnosed long COVID group (τb = –0.178, *p* < 0.001) but not in the self-reported long COVID group (τb = –0.021, *p* = 0.712). Next, we compared the impact of long COVID-19 duration on memory scores between the self-reported long COVID and diagnosed long COVID groups. We combined participants into six-month long COVID-19 duration categories. Fig 4 shows the effect of long COVID duration on memory scores for the self-reported long COVID and diagnosed long COVID groups. A Mann-Whitney U test found no significant difference between the self-reported long COVID and diagnosed long COVID groups for any of the five long COVID duration categories (Bonferroni-corrected for five duration categories).

**Fig 4.**
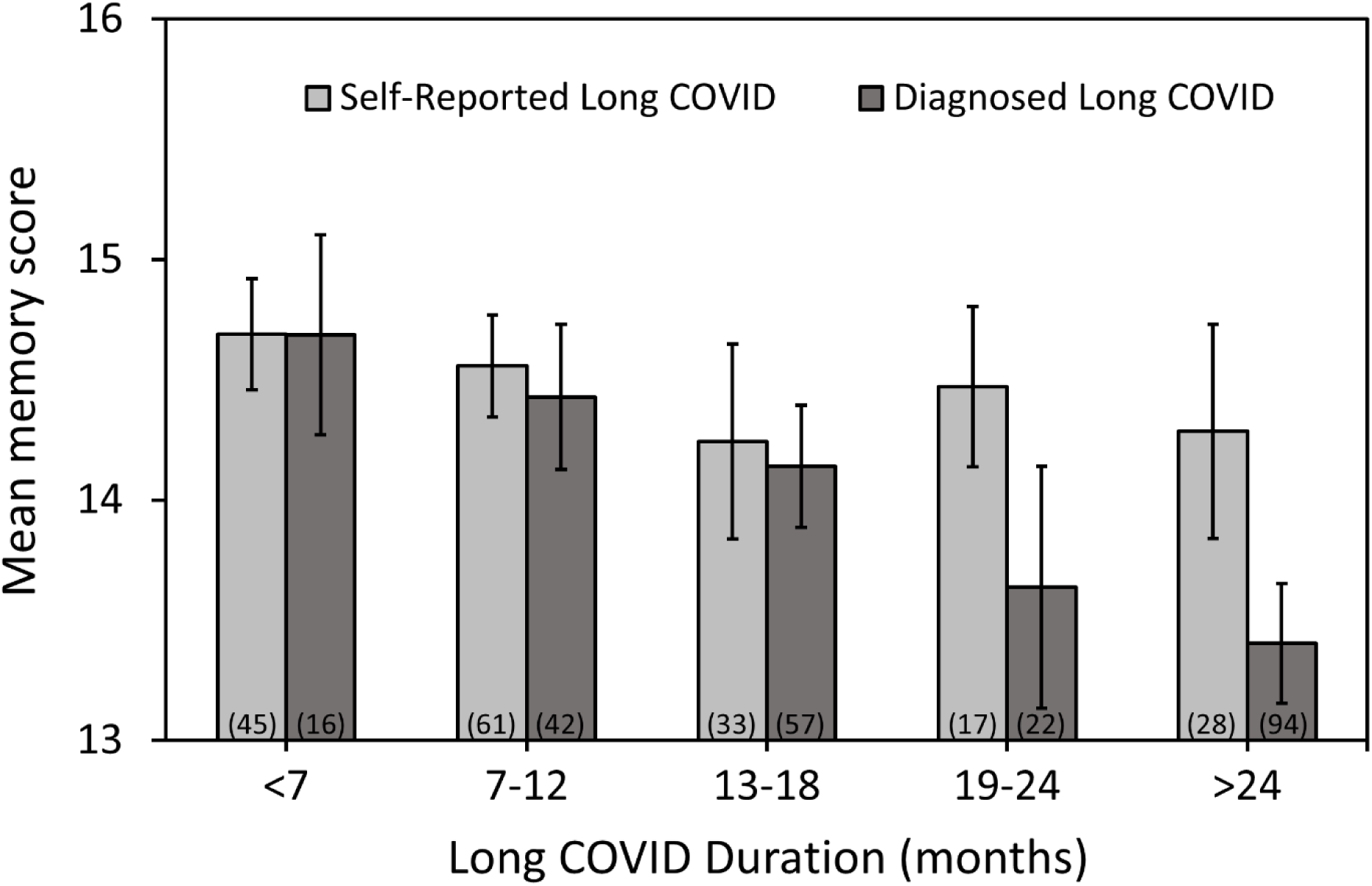
The effect of long COVID duration on objective memory scores in the self-reported and diagnosed long COVID groups. The sample size is given in brackets for each data bar. Note: Each bar represents a different sample of participants (between-subject design). Error bars represent the standard error of the mean.

### COVID-19 variants and working memory

During the COVID-19 pandemic the SARS-CoV-2 virus evolved to produce different variants over time. Consequently, in the current study, those participants who developed long COVID early in the pandemic were likely to have been infected with a different variant than those who were infected later. It has been shown that participants infected when earlier variants were dominant have a higher probability of continuing long COVID symptoms, including cognitive impairment and poor memory (Diexer et al., 2023). Hence, our results indicating lower memory quiz scores with longer COVID duration (Figure 4) could have been influenced by the variant of SARS-CoV-2. When a participant’s SARS-CoV-2 variant is unknown, previous studies have estimated the likely infection strain based on the dominant variant present at a given time period within a given country (Atchison et al., 2023; Diexer et al., 2023; Elliott et al., 2022). In a post-hoc analysis, we divided our long COVID participants from the United Kingdom only into groups according to the dominant variant at the time their long COVID symptoms began. This was achieved by subtracting the number of months since participants’ long COVID symptoms started from the date they completed the online survey and memory quiz. Our variant groups were based on the time periods used previously for the United Kingdom (Atchison et al., 2023; Elliott et al., 2022): 1) wild-type, prior to December 2020; 2) alpha, from the beginning of December 2020 to the end of April 2021; 3) delta, from the beginning of May 2021 to the end of December 2021; 4) omicron, from the beginning of January 2022 onwards.

A Kendall’s tau-b correlation showed a significant relationship between the SARS-CoV-2 variant that was dominant at the time at which long COVID symptoms started and the duration of long COVID symptoms for both the self-reported (τb = −0.740, *p* < 0.001) and diagnosed (τb = −0.869, *p* < 0.001) long COVID groups. Fig 5 illustrates the objective memory scores for each dominant SARS-CoV-2 variant in the self-reported and diagnosed long COVID groups in United Kingdom participants only.

**Fig 5.**
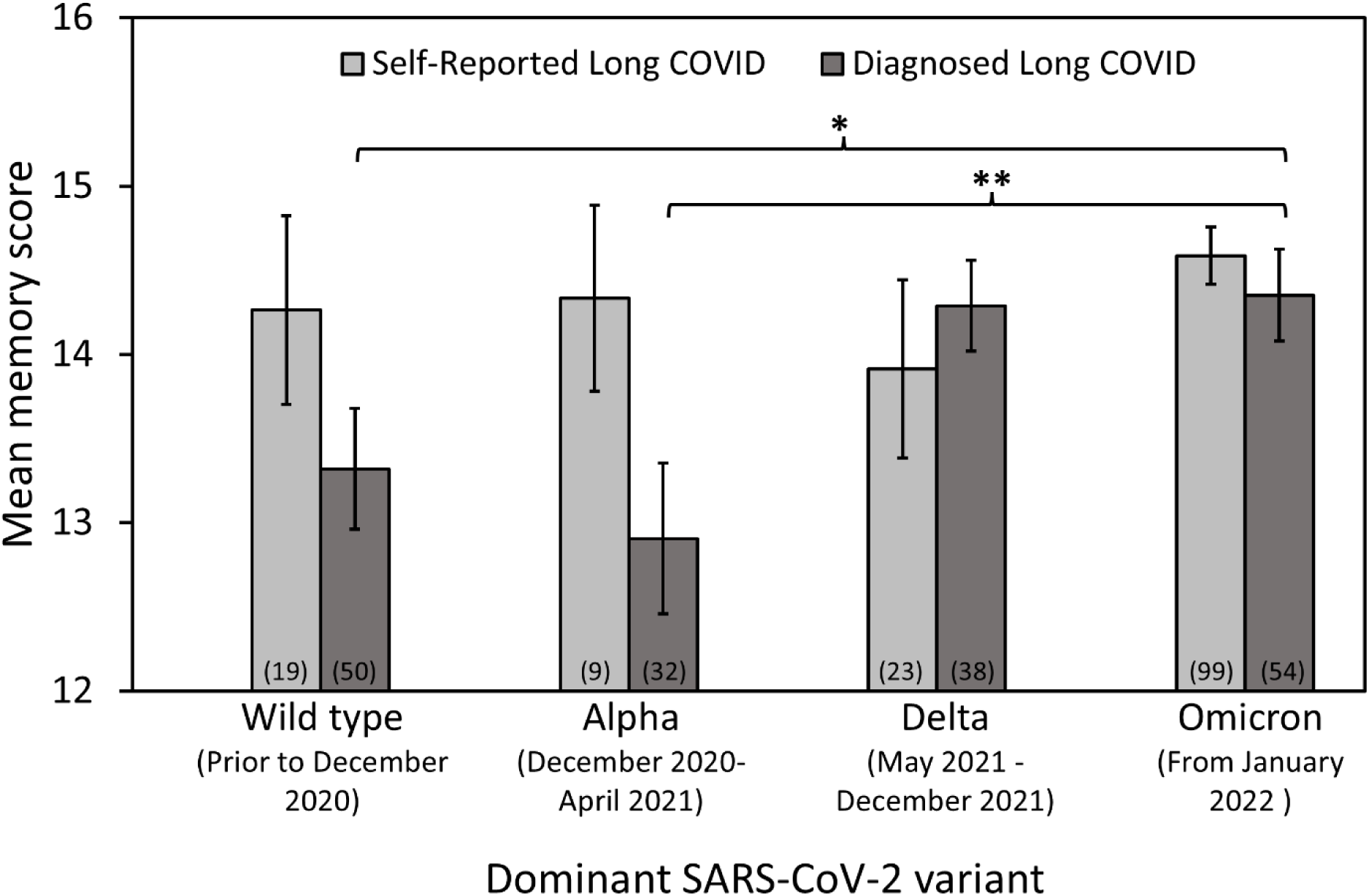
The dominant SARS-CoV-2 variant and objective memory scores in the self-reported and diagnosed long COVID groups in participants only from the United Kingdom. The long COVID participants were divided into four groups (Wild type, Alpha, Delta and Omicron) according to the dominant variant at the time when their long COVID symptoms began. The sample size is given in brackets for each data bar. Error bars represent the standard error of the mean. \**p* < 0.05, \*\**p* < 0.01

For participants with diagnosed long COVID, there was a significant difference in mean memory scores across the four SARS-CoV-2 variants (Kruskal-Wallis test, H(3) = 14.140, *p* = 0.003). For participants with diagnosed long COVID, we found a significantly lower mean memory scores for the wild type (Dunn’s pairwise comparison corrected for multiple comparisons, *p* = 0.033) and alpha (*p* = 0.009) variants compared to the omicron variant. There was no significant difference in mean memory scores across the four SARS-CoV-2 variants for participants with self-reported long COVID (Kruskal-Wallis test, H(3) = 1.236, *p* = 0.744).

### Relationship between ‘*Brain fog’*, ‘*Impact of long COVID symptoms on life’* and objective memory scores

There was a significant negative correlation between the subjective ratings for ‘*Brain fog*’ and objective memory scores across all participants (τb = −0.185, *p* < 0.001). Fig 6a shows the mean subjective ratings for ‘*Brain fog*’ versus objective memory scores for each of the non-long COVID, self-reported long COVID and diagnosed long COVID groups. A Kruskal-Wallis test revealed a significant relationship between long COVID status and ‘*Brain fog*’ ratings [H(2) = 331.90, *p* < 0.001]. Dunn’s pairwise comparisons (with Bonferroni correction for multiple tests) revealed that the subjective ratings of ‘*Brain fog*’ in the self-reported and diagnosed long COVID groups were significantly more severe compared to the non-long COVID group (*p* < 0.001). Moreover, the subjective ratings of ‘*Brain fog*’ in the diagnosed long COVID group were significantly more severe compared to the self-reported long COVID group (*p* = 0.011). There was a significant negative correlation between the subjective ratings for ‘*Impact of long COVID symptoms on life*’ and objective memory scores across all participants with long COVID (τb = −0.169, *p* < 0.001). Fig 6b shows the mean subjective ratings of the ‘*Impact of long COVID symptoms on life*’ versus objective memory scores for the self-reported and diagnosed long COVID groups.

**Fig 6.**
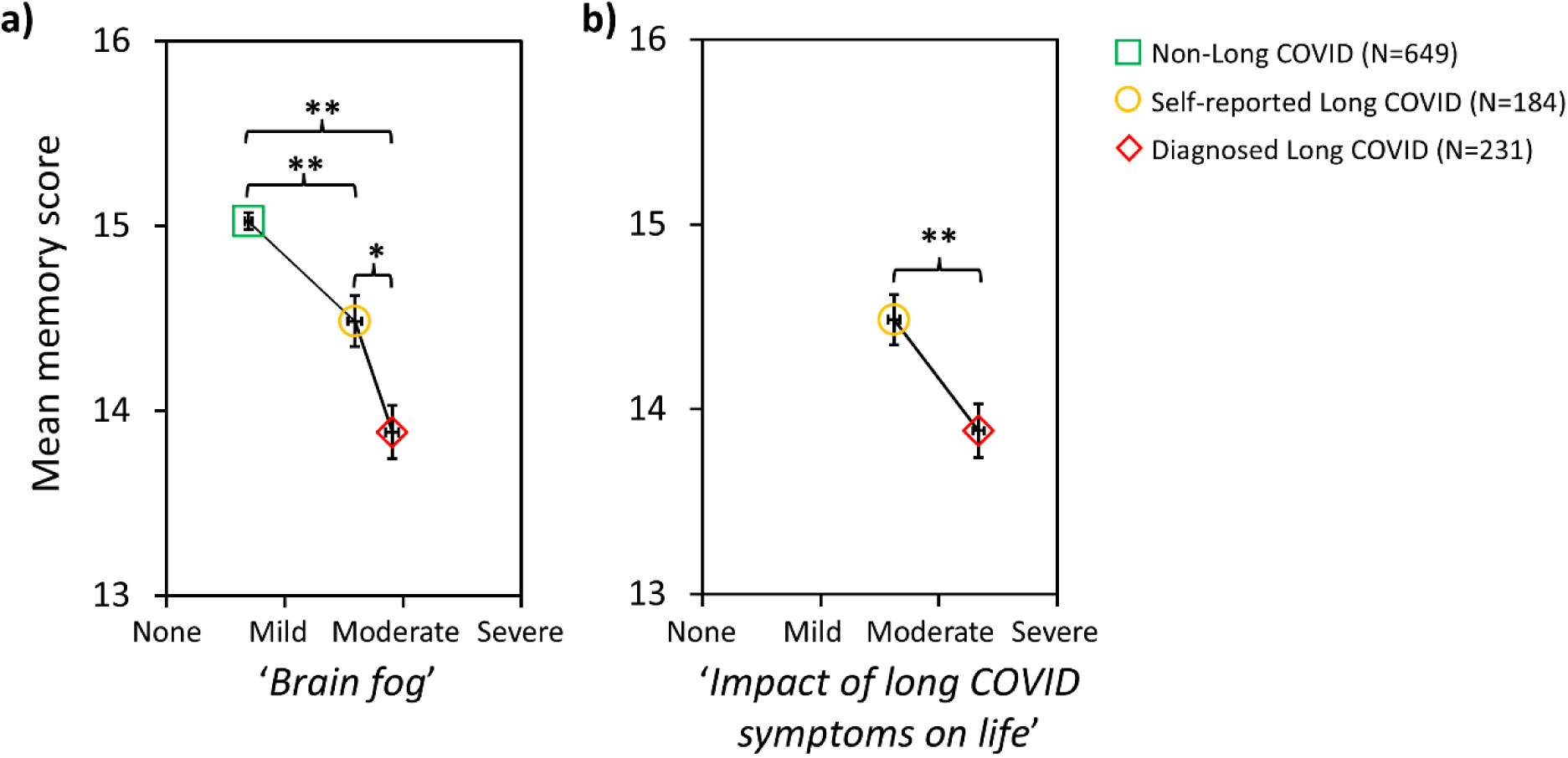
Subjective ratings of a) ‘Brain fog’ and b) ‘Impact of long COVID symptoms on life’ versus memory scores. The vertical error bars represent the standard error of the mean for memory scores, and horizontal error bars indicate the standard error of the mean for ‘Brain fog’ or ‘Impact of long COVID symptoms on life’ ratings. Statistical significance values relate to ratings for ‘Brain fog’, and for ‘Impact of long COVID symptoms on life’ \**p* < 0.01, \*\**p* < 0.001.

The subjective ratings of ‘*Impact of long COVID symptoms on life*’ in the diagnosed long COVID group were significantly worse compared to the self-reported long COVID group (Mann-Whitney U test = 10372.00, Z = −9.61, *p* < 0.001, r = 0.47). There was a significant correlation between the subjective ratings for ‘*Brain fog*’ and ‘*Impact of long COVID symptoms on life*’ in the long COVID (self-reported and diagnosed) group (τb = 0.405, *p* < 0.001).

## Discussion

The main finding in our study was that long COVID negatively impacts upon working memory function. The most important factor affecting working memory scores was long COVID status (diagnosed long COVID, self-reported long COVID and non-long COVID) followed by age and number of COVID-19 infections. Using the same single rapid online working memory test as in our previous study (Baseler et al., 2022), we found that the objective memory scores were significantly reduced in the long COVID groups (diagnosed with long COVID or self-reported having long COVID) compared to the non-long COVID group. These results are in line with findings from other studies showing memory deficits in individuals with continuing COVID-19 symptoms (Bertuccelli et al., 2022; Graham et al., 2021; Guo et al., 2022; Miskowiak et al., 2021; Miskowiak et al., 2023; Zhao et al., 2023). We found that the mean memory score for those diagnosed with long COVID (13.88 ± 0.15) was lower than for those in our previous study who had tested positive for COVID-19 but had not been hospitalized (14.73 ± 0.06) and more comparable to people who had been hospitalized with COVID-19 (13.46 ± 0.41) (Baseler et al., 2022). Together, these findings indicate that working memory is particularly impaired in those diagnosed with long COVID.

Our results provide evidence that working memory deficits may be linked to ‘brain fog’, one of the most commonly reported symptoms in those with long COVID (Aghajani Mir, 2023; Kalak et al., 2022; Millet et al., 2022; Premraj et al., 2022). We found that for all participants, objective memory scores were negatively correlated with increasing levels of brain fog, such that those with lower memory scores reported the most severe brain fog. The mean subjective ratings of brain fog were most severe for the diagnosed long COVID group followed by the self-reported long COVID group and were mildest in those who did not have long COVID. Our results align with those of a previous study on long COVID, where memory impairments were self-reported in 66.2% of participants with brain fog compared to 13.5% without brain fog (Jennings et al., 2022). The mean subjective ratings of the ‘*Impact of long COVID symptoms on life*’ were also most severe for the diagnosed long COVID group with lower memory scores compared to the self-reported long COVID group who had higher memory scores. Our results suggest that poorer working memory may be one of the many symptoms that could contribute to the negative impact of long COVID on daily life. This is further supported by our finding that subjective ratings for ‘*Brain fog*’ and ‘*Impact of long COVID symptoms on life*’ were highly correlated. Previous studies have also shown that aspects related to the lived experience of long COVID can significantly reduce quality of life, including persistent physical symptoms, mental health problems, ability to cope, reduced functional status and employment issues (Aiyegbusi et al., 2021; Graham et al., 2021; Kennelly et al., 2023; Kim et al., 2023; Natarajan et al., 2023; Ziauddeen et al., 2022).

Previous studies on cognitive function in long COVID have typically classified participants into a long COVID group based on persistent symptomology rather than clinical diagnosis by a medical professional (Bungenberg et al., 2022; Guo et al., 2022; Lauria et al., 2023; Thronicke et al., 2022). One of the strengths of our study is that we included a group of long COVID participants who were clinically diagnosed. Medical professionals will diagnose patients with long COVID based on multiple sources of evidence, including case history, signs and symptoms, physical assessments/clinical tests, and may be supported by members of a multi-disciplinary team. In our study, we added a separate self-reported long COVID group to represent those who may have long COVID but were not clinically diagnosed.

Furthermore, to guide participants who think they may have long COVID we provided in our survey an explicit definition from NHS England of long COVID, namely, ‘*Signs and symptoms that develop during or after COVID-19 and continue for more than 12 weeks and are not explained by an alternative diagnosis*.’

We found that the objective memory scores decreased significantly with age in the non-long COVID control group. This finding aligns with the age-related effects observed in the control group of our previous study, which utilized the same working memory quiz (Baseler et al., 2022). It is also consistent with other research demonstrating that working memory declines as age increases (Brockmole & Logie, 2013; Park et al., 2002). Importantly, the reduction of working memory scores with age was greater in the group diagnosed with long COVID compared to the non-long COVID group. Specifically, long COVID had a greater impact on memory scores for the older groups categories (35 years old and above) compared to the younger age categories (<35 years old) where there was no significant difference in memory score between the diagnosed and non-long COVID groups. Our data indicate that working memory is impacted most by long COVID in adults aged 35 years and above. However, this interpretation must be contextualized by considering sample size, since the number of participants diagnosed with long COVID was lowest in the youngest age grouping (16-24 years) and higher in the older age groups. Our distribution reflects the prevalence of long COVID across age groups found in the general population (ONS, 2023). Nevertheless, our finding that memory problems in long COVID are more pronounced in older age groups aligns with the broader observation that older individuals experience greater memory impairments post-COVID-19 (Baseler et al., 2022; Blomberg et al., 2021; Corbett et al., 2023; Cui et al., 2024; Finamore et al., 2024; Nalbandian et al., 2021; Stavem et al., 2022).

A study examining SARS-CoV-2 reinfection found that having two or more infections increased the risk of persistent symptoms in the post-acute phase (up to six months after infection) including neurological sequelae (Bowe et al., 2022). In our current investigation we found that objective memory scores decreased with increasing number of COVID-19 infections but only in the group diagnosed with long COVID. This finding supports the viewpoint that COVID-19 reinfections increase the severity of cognitive dysfunction in people with long COVID. There were a few participants who reported they had not had COVID-19 infection, but self-reported as having long COVID (N=5) or were diagnosed with long COVID (N=1). There are several possible reasons which could account for this, including the absence of COVID testing at the time of infection, being asymptomatic to the COVID-19 infection (Gao et al., 2021), a side effect of the COVID-19 vaccine (Finterer & Scorza, 2022), or simply an error when completing the questionnaire.

In our cross-sectional study, we found that memory scores decreased as the duration of long COVID symptoms persisted in those diagnosed with long COVID. This suggests that working memory function progressively deteriorates following COVID-19 infection in individuals with long COVID. However, the effects of long COVID duration on working memory scores could be confounded with the effects of the different SARS-CoV-2 variants that infected participants. A previous investigation has indicated that variant type affects the severity of cognitive deficits, with earlier variants such as wild-type and alpha causing greater deficits than more recent delta and omicron variants (Hampshire et al., 2024). In a post-hoc analysis, we divided participants from the United Kingdom with long COVID into four variant groups (wild-type, alpha, delta and omicron) based on dominant SARS-CoV-2 strains according to the time periods defined in previous studies (Atchison et al., 2023; Elliott et al., 2022). We found that the duration of long COVID symptoms was negatively correlated with the date order of dominant strains, 1) wild-type, 2) alpha, 3) delta and 4) omicron, in both self-reported and diagnosed long COVID groups. This suggests that individuals with longer duration symptoms were more likely to be infected with earlier variants of the virus. In the diagnosed long COVID group, those whose symptoms began when the wild-type and alpha variants were prevalent had lower memory scores compared to the later omicron variant. Although our results suggest that having long COVID for a protracted time is detrimental to working memory, this interpretation must be considered within the context of the prevalent SARS-CoV-2 strain. To further address the impact of long COVID duration on memory without the confound of SARS-CoV-2 variants, a future longitudinal approach is suggested where individuals’ memory scores are assessed at multiple timepoints. Such longitudinal studies could also correlate individuals’ memory scores with their health-related clinical data, including self-reported brain fog severity, to contextualize long-term trends of cognitive recovery or decline in long COVID. Since our interpretation of the effects of COVID-19 variant on memory scores was limited by assuming a dominant strain based on date alone, future studies could test individuals for variant type to extract the separate contribution of variant from duration.

We found that the group diagnosed with long COVID had lower scores compared to those who self-reported having long COVID indicating that working memory deficits were greater in those diagnosed by a medical professional. Moreover, in the self-reported long COVID group, memory scores did not decrease significantly with age, number of COVID-19 infections, long COVID duration or probable SARS-CoV-2 variant. It is possible that participants were more likely to seek help and therefore be diagnosed with long COVID by medical professionals if they had more severe long COVID symptoms, including brain fog, memory problems and overall impact on their lives. It is possible that the self-reported group may have had higher memory scores than the diagnosed group because they had less severe symptoms and felt less of a need to consult a medical professional. Our study supports this hypothesis, as we found that the diagnosed long COVID group reported that they had more severe brain fog and that their symptoms had a greater impact on their life. A previous study has shown that more severe COVID-19 symptoms are linked to poorer cognitive function including memory deficits (Guo et al., 2022). In addition, some participants in the self-reported long COVID group may have incorrectly diagnosed themselves based on their interpretation of the NHS single sentence definition of long COVID. Consequently, it is possible that these participants did not actually have long COVID and that their memory was less affected resulting in higher memory scores compared to the diagnosed group. The misdiagnosis of long COVID could affect the interpretation of results from prior studies that relied on self-reported data to classify the condition and may have led to an underestimation of the impact of long COVID on memory.

Several mechanisms have been proposed to explain the impact of long COVID on cognition including memory function. These include pathophysiological mechanisms that could impede brain function such as neuroinflammation, damage to neurons and astrocytes, vascular alterations, neurotropic effects, dysautonomia, hypoxia, blood coagulation, and metabolic changes (Bauer et al., 2022; Davis et al., 2023; Moller et al., 2023; Nalbandian et al., 2021; Natarajan et al., 2023; Plantone et al., 2024; Turner et al., 2023). In addition, long COVID could affect psychological well-being, leading to conditions such as anxiety, depression, post-traumatic stress disorder, or sleep disturbances which in turn could contribute to memory deficits (Aiyegbusi et al., 2021; Davis et al., 2023; Kennelly et al., 2023; Natarajan et al., 2023; Tedjasukmana et al., 2022).

Since our main result demonstrates that long COVID can lead to problems with working memory, healthcare providers could incorporate a test which specifically assesses working memory when evaluating long COVID and to monitor changes over time following interventions. A number of rehabilitation approaches have been proposed to treat long COVID cognitive symptoms (with mixed effectiveness) including cognitive training, electromagnetic brain stimulation, psychosocial, pharmaceutical, natural supplements, psychoeducation/compensatory skills training, telerehabilitation and physical rehabilitation such as hyperbaric oxygen therapy (Gorenshtein et al., 2024; Hawke et al., 2024; Moller et al., 2023; Salawu et al., 2020). A personalized clinical treatment plan that considers working memory deficits may help address both the cognitive as well as physical symptoms of long COVID.

### Study limitations

While the quiz was designed based on established working memory paradigms and has previously demonstrated sensitivity to COVID-related cognitive differences (Baseler et al., 2022), we cannot make definitive claims about its reliability, validity, or specificity. However, our approach offers a pragmatic solution for screening larger populations for working memory difficulties in long COVID, which may then inform more detailed clinical evaluation. Our working memory quiz is currently being applied along with several standard neuropsychological tests including the Montreal Cognitive Assessment (MoCA) and the Modified Mini-Mental State Exam (3MSE) in a study evaluating cognitive function in patients with kidney disease undergoing haemodialysis (Aksoy et al., 2024). Future research should aim to validate this brief online measure against established clinical neuropsychological tests to determine its psychometric properties, including test-retest reliability, construct validity, discriminant validity, and sensitivity to change over time. Prior to starting the memory quiz, the survey reminded participants of the importance of the study and to undertake the quiz in an environment without distractions.

However, we acknowledge the inherent limitations of unsupervised online testing, including variable levels of distraction and compliance, which could affect performance of any participant regardless of long COVID status. Nonetheless, online testing offers significant advantages in terms of accessibility, particularly important for reaching individuals with mobility limitations including those with long COVID. The ecological validity of testing participants in more natural environments may, however, better reflect real-world cognitive functioning than laboratory-based assessments.

Our use of convenience sampling through social media, support groups, and professional networks may have introduced bias. For example, individuals engaged in long COVID support groups may be those who experience more persistent or severe symptoms, which could lead to an overestimation of cognitive difficulties.

However, there may have also been an underrepresentation of those too unwell to complete the study, which could contribute to an underestimation of effects in the most affected group. The majority of our sample was based in the United Kingdom (83%), which may limit the generalizability of the findings worldwide. Capturing demographics and epidemiology of long COVID across countries is challenging because studies across the world often use different sampling methods, inclusion criteria, and methodologies, making it difficult to consistently compare and combine datasets. However, the distribution of ages and gender in our sample are in line with the demographics published in a comprehensive review of long COVID which includes data from the Office of National Statistics in the United Kingdom (Greenhalgh et al., 2024).

Although an *a priori* power analysis was not conducted before data collection, with our final sample size of 1064 participants (415 with long COVID and 649 controls) and an effect size of Hedges’ g = 0.552, the calculated statistical power was > 99%. Moreover, after correction for multiple comparisons, all statistically significant differences resulted in *p*-values < 0.005. Together, this indicates that our study is sufficiently powered to detect differences in working memory scores using our single rapid online test.

Our study intentionally did not require positive COVID-19 test confirmation, aligning with NHS England and WHO clinical definitions of long COVID, which do not mention the requirement for a positive COVID-19 test. This pragmatic approach recognizes the limited testing availability during the early period of the pandemic. PCR testing in the UK was initially restricted to hospital settings (February 2020) and only gradually expanded to the general public, with widespread home testing (lateral flow test) available from April 2021. Among our participants reporting long COVID symptoms, only six indicated they had never had COVID-19 infection, a minimal proportion that is unlikely to affect our findings. However, we acknowledge that retrospective self-reporting of COVID-19 status has limitations, as participants may not, for example, recall testing status accurately or may have had asymptomatic infections.

The purpose of the study was to compare working memory function in those with long COVID with those without long COVID, and not to determine the specific role of all the various factors involved. We operated under the assumption that participants with pre-existing cognitive impairments or other conditions affecting working memory were equally distributed across both non-long COVID and long COVID cohorts, but this may not have been the case. In our study, we investigated several factors that could have contributed to memory function in long COVID (age, number of COVID-19 infections, long COVID duration, and brain fog and symptom severity). Long COVID is a complex multi-factorial condition that affects individuals differently.

Individuals with long COVID might have a higher prevalence of factors affecting working memory, including mental and neurological conditions (such as depression or anxiety), fatigue, medications, or differences in education levels, or the potential interactions between these factors and others measured in our study, such as brain fog. Based on our results, future studies could be designed to disentangle the specific contributions of these and other factors on memory function in long COVID.

## Conclusions

This study demonstrates that long COVID, particularly in clinically diagnosed cases, is associated with significant impairments in working memory. These memory impairments are exacerbated by factors such as age and the number of COVID-19 infections. Additionally, individuals reporting more severe brain fog and greater overall life impact from long COVID exhibited lower memory scores. These findings underscore the importance of implementing targeted interventions for individuals with long COVID, with a focus on addressing cognitive deficits and improving quality of life.

## Materials and methods

### Ethics and participant recruitment

The study was undertaken in accordance with the Declaration of Helsinki (World Medical, 2013). Ethical approval was given by the Hull York Medical School Ethics Committee (Reference 22-23 41). The Long COVID Online Rapid Objective Neuro-Memory Assessment (L-CORONA) was made publicly available, and data collected from 16/03/2023 to 31/12/2023. The link to the online survey and memory quiz was disseminated through multiple channels, including: 1) Direct communication through professional networks and healthcare contacts, 2) University and healthcare organization intranets, 3) Social media platforms, including long COVID support groups, 4) Advertisements posted in public places and community settings, and 5) Word-of-mouth referrals from participants. This convenience sampling approach was chosen to maximize participation and reach individuals both with and without long COVID. Participants aged 16 years old and over were invited to complete the survey and memory quiz regardless of whether they had long COVID. After providing information at the beginning of the survey about the study, written informed consent was obtained by asking participants, ‘*Do you consent to take part?*’, and only those who answered ‘*Yes*’ were allowed to proceed. The L-CORONA survey and memory quiz complied with General Data Protection Regulations (GDPR), was fully anonymous, and no personal contact details were acquired. The Qualtrics platform (University of York license) collected and stored the encrypted data, and access was restricted to the study investigators only. Since only anonymized responses were captured, any data made publicly available contains no personal, identifiable information. No compensation was provided to participants for completing the assessment.

### Long COVID definitions

Some studies reporting the impact of COVID-19 on cognitive function do not provide an explicit definition of long COVID but instead have inferred long COVID status based upon ongoing symptomology (Bungenberg et al., 2022; Guo et al., 2022; Lauria et al., 2023; Thronicke et al., 2022). The NHS England’s definition of post COVID-19 syndrome, which is based on the National Institute for Health and Care Excellence definition (NICE, 2020), is ‘*…signs and symptoms that develop during or after COVID-19 and continue for more than 12 weeks and are not explained by an alternative diagnosis*’ (NHS, 2021). This definition is broadly consistent with that of the World Health Organization which defines post COVID-19 condition (long COVID) as, ‘*…the continuation or development of new symptoms 3 months after the initial SARS-CoV-2 infection, with these symptoms lasting for at least 2 months with no other explanation*’(WHO, 2022). In our current study, we adopted the NHS England definition of long COVID with a questionnaire designed to exclude people who had symptoms less than 12 weeks after being infected with COVID-19. In addition, we designed our study to compare people diagnosed with long COVID by a medical professional with those who self-reported having long COVID. This approach balances the challenges of obtaining formal clinical diagnoses for all long COVID participants by including both clinically validated and self-reported cases. It acknowledges that many individuals may meet symptom criteria without formal diagnosis due to barriers including healthcare access, financial constraints, geographic isolation, or healthcare providers not recognizing or validating reported long COVID symptoms. By incorporating all participants who meet defined criteria, this methodology mitigates selection bias while enabling meaningful comparison between diagnosed and self-reported groups.

### Long COVID survey and memory quiz structure

The long COVID assessment was similar in structure to our previous COVID-19 online survey and memory quiz (Baseler et al., 2022). This memory quiz was designed based on established principles of working memory assessment, incorporating a visual recognition paradigm that tests short-term storage and retrieval of visual information across different semantic categories. In our previous study, this quiz successfully discriminated between individuals with and without COVID-19 infection, was sensitive to infection severity (hospitalized versus non-hospitalized) and correlated with participants’ self-reported assessment of their cognitive function and symptom severity, all of which support the construct validity of our working memory quiz (Baseler et al., 2022). The quiz was intentionally designed to be brief and engaging to maximize completion rates and minimize participant fatigue, which is particularly important when testing individuals with long COVID who frequently report fatigue as a primary symptom. The memory task’s design prioritizes ecological validity and accessibility over the laboratory-controlled conditions typical of traditional cognitive assessments, representing a trade-off that allowed us to collect data from a larger and more diverse sample.

The Qualtrics platform (University of York license) was used to develop the online survey and memory quiz. Participants accessed the long COVID assessment using a Uniform Resource Locator (URL) or a quick response (QR) code, which made it available on smartphones, tablets, laptops and PCs with internet access. No time limit was imposed upon participants to complete the long COVID assessment. Screenshots of some survey questions and a sample of the memory quiz are illustrated in Fig 7.

**Fig 7.**
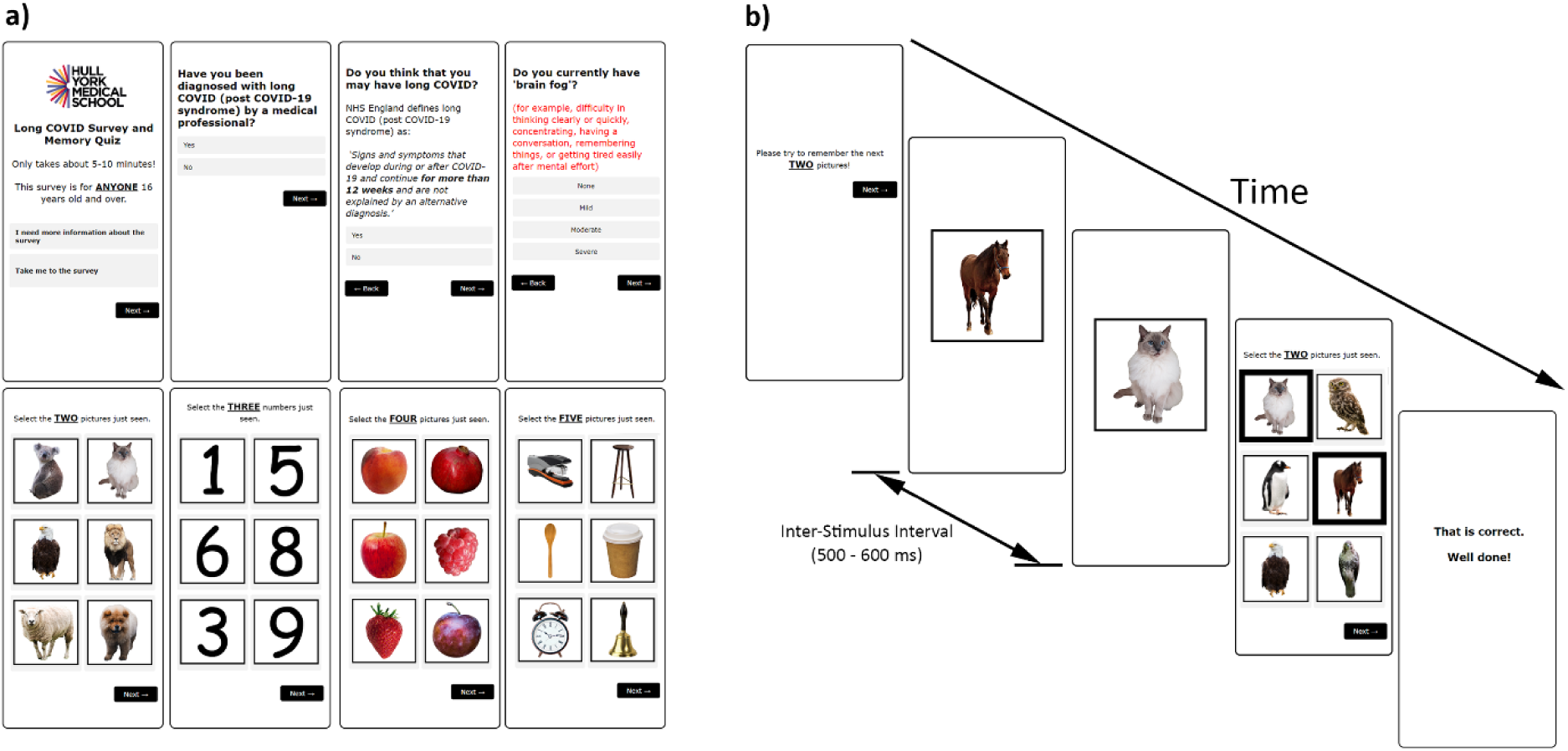
The ‘Long COVID Survey and Memory Quiz’ presented on a smartphone. **a)** Example questions from the survey, and examples of the animal, number, fruit and object categories shown from the memory quiz. **b)** A series of screenshots representing a single trial from the working memory quiz.

The survey component consisted of a series of ‘*Yes/No*’ questions asking participants about their long COVID status. First, we asked, ‘*Have you been diagnosed with long COVID (post-COVID-19 syndrome) by a medical professional?*’. If a participant answered ‘*Yes*’, they were designated into the diagnosed long COVID group. Those answering ‘*No*’ were then asked, ‘*Do you think that you may have long COVID?*’, and we included the NHS England definition of long COVID (post COVID-19 syndrome): ‘*Signs and symptoms that develop during or after COVID-19 and continue for more than 12 weeks and are not explained by an alternative diagnosis*.’ (NHS, 2021). Those that answered ‘*Yes*’ to this question were designated as the self-reported long COVID group. Those who answered ‘*No*’ to both questions were designated as the non-long COVID group. If they were diagnosed with long COVID or self-reported having long COVID, they were asked how many months it had been since their long COVID symptoms started (using a drop-down menu, from less than 3 months to 60 months). We next asked long COVID participants to subjectively rate the overall life impact of their long COVID symptoms (using *‘None’, ‘Mild’, ‘Moderate’ or ‘Severe’*). All participants (regardless of long COVID status) were asked whether they had been infected with COVID-19 in the last 12 weeks (‘*Yes/No*’), and how many times they had been infected with COVID-19 (‘*Not had COVID-19*’, ‘*One*’, ‘*Two*’, ‘*Three*’, or ‘*Four or more times*’). All participants were also asked to subjectively rate their level of ‘*brain fog*’ (using *‘None’, ‘Mild’, ‘Moderate’ or ‘Severe’*). The following definition of ‘*brain fog*’ was provided to aid participants: ‘*for example, difficulty in thinking clearly or quickly, concentrating, having a conversation, remembering things, or getting tired easily after mental effort’.* This definition of brain fog was based on our previous findings showing that these COVID-19 cognitive symptoms were highly correlated with one another (Baseler et al., 2022). Finally, participants were asked to select their age range (16-17, 18-24, 25-34, 35-44, 45-54, 55-64, 65-74, 75-84 and 85+), gender, and country of current residence. Following completion of these survey questions, participants next completed the objective working memory quiz. To increase levels of concentration and compliance, the participants were reminded of the importance of the study (*Time for a Memory Quiz! Good luck! Your answers will help us to understand the effect of long COVID on memory.*’) and instructed to ‘*Please complete the quiz in a QUIET place WITHOUT DISTRACTIONS.*’ The memory quiz consisted of the same visual working memory/recall task that we utilized previously to investigate the impact of COVID-19 on working memory (Baseler et al., 2022). In brief, the working memory quiz employed aspects of gamification such that the task was relatively easy to understand with familiar and engaging images. Participants progressed through the quiz with increasing difficulty levels and motivational feedback was given for responses. At the end of the quiz, participants were notified of their overall score along with the scores for the sub-categories. Four image categories were displayed to participants consisting of animals, numbers, fruits, and objects (Fig 7a). The stimulus set comprised of 55 unique images: animals, fruits, objects (15 unique images in each of these categories), and ten single-digit whole numbers (0 to 9). For each image category, two, three, four or five different random images were shown consecutively. Each image was presented for 500ms and the interstimulus interval was randomly varied between 500-600ms (Fig 7b). A grid of six images was presented, and participants were instructed to select the images (two, three, four or five) viewed prior. Participant responses were not time restricted.

### Data analysis

Based on our inclusion criteria, participants under the age of 16 were excluded (N=3). Participants that had COVID-19 infections within the last 12 weeks were also excluded from the analysis (N=146), as a recent COVID-19 infection is known to affect working memory (Baseler et al., 2022). In addition, we excluded one participant who reported having long COVID for 49 months before completing the survey/memory quiz on 21^st^ March 2023. This meant that their long-COVID symptoms would have started on the 21^st^ of February 2019, a time before COVID-19 was identified.

A score of 1 was given for each correct trial in the memory quiz, yielding a possible maximum objective memory score of 16. Statistical Package for the Social Sciences (SPSS Version 29.0.1.0, IBM Corporation, Armonk, N.Y., USA) was utilized to perform statistical analyses. To minimize the risk of Type I errors from multiple comparisons while maintaining sensitivity to potential effects, we employed a hierarchical analytical approach. As working memory scores were not normally distributed, we first used categorical regression (Optimal scaling-CATREG, SPSS) to identify which independent variables significantly contributed to variance in memory scores. In the first categorical regression, we assessed the contributions of the following independent variables: long COVID status (three groups: non-long COVID, diagnosed long COVID and self-reported long COVID), age (nine groups: 16-17, 18-24, 25-34, 35-44, 45-54, 55-64, 65-74, 75-84 and 85+), number of COVID-19 infections (five groups: none, 1, 2, 3, and 4 or more) and gender (three groups: female, male, other/prefer not to say). A second categorical regression was performed using the long COVID groups only with the following independent variables: long COVID status (two groups: diagnosed long COVID and self-reported long COVID), age, number of COVID-19 infections, gender, and long COVID duration in months (from less than 3 months up to 60 months). Next, we investigated the effect of the significant individual variables arising from the categorical regression analysis on objective memory scores, using non-parametric statistics: Mann-Whitney U test and Kruskal-Wallis test with Dunn’s pairwise comparisons (Bonferroni corrected). We evaluated relationships between pairs of variables using the Kendall’s tau-b correlation coefficients. All testing was two-tailed, and we set statistical significance levels to *p* values less than 0.05.

## Data Availability

All relevant data will be made available in Supporting Information files following manuscript acceptance for publication in a journal.

## Acknowledgments

We are grateful to Namitha Thottiyil for assistance in participant recruitment.

## Author Contributions

**Conceptualization:** Aziz U. R. Asghar, Heidi A. Baseler, Abayomi Salawu, Murat Aksoy.

**Data curation:** Murat Aksoy, Aziz U. R. Asghar.

**Formal analysis:** Aziz U. R. Asghar, Hong Kiu Yuen, Heidi A. Baseler.

**Investigation:** Aziz U. R. Asghar, Hong Kiu Yuen, Murat Aksoy, Heidi A. Baseler

**Methodology**: Aziz U. R. Asghar, Hong Kiu Yuen, Murat Aksoy, Abayomi Salawu, Heidi A. Baseler.

**Project administration**, Aziz U. R. Asghar, Heidi A. Baseler.

**Resources:** Aziz U. R. Asghar, Murat Aksoy, Heidi A. Baseler.

**Supervision**: Aziz U. R. Asghar, Abayomi Salawu, Heidi A. Baseler.

**Visualization:** Aziz U. R. Asghar, Heidi A. Baseler.

**Writing – original draft:** Aziz U. R. Asghar, Heidi A. Baseler.

**Writing – review & editing:** Aziz U. R. Asghar, Hong Kiu Yuen, Murat Aksoy, Abayomi Salawu, Heidi A. Baseler.

